# Centre-specific treatment and outcomes of patients with brain metastases treated with whole brain or stereotactic radiation therapy: an Ontario-based retrospective cohort study

**DOI:** 10.1101/2025.10.10.25337749

**Authors:** Andrew Youssef, Arjun Sahgal, Daniel Felsky, Aisha Lofters, Sunit Das

**Affiliations:** Institute for Medical Sciences, University of Toronto, Toronto, ON; Department of Radiation Oncology, Sunnybrook Hospital, University of Toronto, Toronto, ON; Department of Psychiatry, University of Toronto, Toronto, ON; Krembil Centre for Neuroinformatics, Centre for Addiction and Mental Health, Toronto, ON; Institute for Clinical Evaluative Sciences, Toronto, ON; Dalla Lana School of Public Health, University of Toronto, Toronto, ON; Department of Family and Community Medicine, Women’s College Hospital, University of Toronto, Toronto, ON; Division of Neurosurgery, St. Michael’s Hospital, University of Toronto, Toronto, ON

**Keywords:** Cancer, Brain Metastases, Whole Brain Radiation Therapy, Stereotactic Radiation Therapy

## Abstract

**Background:** Whole brain radiation therapy (WBRT) and stereotactic radiation therapy (SRT) are the two most common treatments for patients with intracranial metastatic disease (IMD). In Ontario, Canada, 4 of the 18 (22%) regional cancer centres (RCC) are fitted with dedicated cranial SRT program, which we refer to as SRT-dedicated program centres, while the remaining RCCs deliver SRT but lack a dedicated cranial SRT program. We hypothesized that SRT usage would be higher and patient outcomes better in RCCs with vs. those without an SRT-dedicated program.

**Methods:** Using administrative data housed at the Institute for Clinical Evaluative Sciences (IC/ES), we identified patients who had a primary cancer diagnosis and received cranial RT between April 1, 2010, and August 31, 2023. Treatment centres were grouped based on the presence or absence of an SRT-dedicated program. Within these groupings, hazard ratios (HR) for overall survival (OS) were compared. Additional comparisons based on the movement of patients across centres were performed.

**Results:** The median OS of the cohort was 6.05 months. Significant differences in treatment and patient characteristics were observed according to RT modality used at RCCs with SRT-dedicated programs vs. those without, including the ratio of WBRT vs SRT (OR = 2.8, 95% CI 2.4-3.2, p < 0.0001), patient age (*X*^2^ = 38.2, df = 7, p < 0.0001), stage at diagnosis (*X*^2^=50.1, df = 4, p < 0.0001), and number of brain metastases treated (OR = 29.9, 95% CI 14.5-76.9, p < 0.0001). The median OS for patients treated with intracranial RT (WBRT or SRT) at centres with vs. those without a dedicated SRT programs was 7.20 vs 5.03 months (HR = 0.81, p < 0.0001), respectively. OS for patients who were treated with SRT at centres with vs. without a dedicated SRT program was 10.15 vs 7.89 months (HR = 0.88, p = 0.0275), respectively. No significant difference in the median OS for patients treated with WBRT at centres with or without a dedicated SRT program was observed (2.96 vs 3.09 months, HR = 1.08, p = 1.0, respectively). Mean OS for patients who travelled to a centre with a dedicated SRT program for intracranial RT after initial systemic therapy at a non-SRT-dedicated centre was associated with an positive impact on OS, compared to OS for patients who received systemic therapy and RT at a non-SRT-dedicated centre (14.0 vs 7.3 months, t = -3.9, p < 0.001).

**Conclusion:** Our study found greater utilization of SRT and significantly longer OS for patients with IMD treated in Ontario RCCs with an SRT-dedicated program. OS was not negatively impacted for patients who travelled to a centre with a dedicated SRT program for radiation therapy.

## Introduction

Intracranial metastatic disease (IMD) occurs in roughly 2% of all cancer patients, accounts for about 12% of all metastatic disease, and poses a substantial negative effect on patient survival and quality of life.^1^ Radiation therapy (RT) is a cornerstone of IMD treatment with increasing utilization of focal treatments to brain metastases with stereotactic radiation as opposed to whole brain radiation (WBRT).^2^

Stereotactic radiosurgery (SRS) refers to radiation therapy delivered in a single fraction. Hypofractionated stereotactic radiation therapy (HSRT) refers to stereotactic delivery of 2 to 5 fractions delivered radiation with a minimum dose per fraction of 5Gy. Henceforth, both SRS and HSRT will be referred to as stereotactic radiation therapy (SRT).^3^ Randomized control trials support the use of SRT instead of WBRT in patients with four or fewer brain metastases (BrM), with increasing evidence supporting better neurocognitive outcomes for patients with up to 10-20 BrM ^2,4,5,6^. Current guidelines recommend SRT in patients with low extra-cranial tumour burden and a good expected prognosis; there is increasing practice of offering SRT to these patients regardless of the number of BrM as long as technically feasible.^2^

In Ontario, Canada, cancer care is delivered at regional cancer centres (RCC), which have capacity to deliver both RT and systemic therapy. RCCs have partnerships with centres in more remote areas, which are able to deliver systemic therapy but are not equipped to deliver RT (non-RT centres). While WBRT and SRT capabilities are available in 17 of the 18 RCC in Ontario, only four RCCs have a dedicated intra-cranial RT apparatus (i.e. Gamma Knife or Cyberknife) and on-site neurosurgical services, while the remaining 14 RCCs utilize conventional image-guided LINAC-based SRT, with or without on-site neurosurgical services. We refer to the former as SRT-dedicated centres and latter as non-SRT-dedicated centres.

We hypothesized that SRT-dedicated centres would be more likely to utilize SRT for the treatment of patients with IMD, and for patients with more BrM, than non-SRT-dedicated centres, and that variations in treatment approach will be associated with differences in patient outcomes. We also aimed to determine if there are negative impacts on patients who travel for SRT while undergoing systemic therapy care in non-RT centres.

## Methods

### Population & study design

This study was conducted as a retrospective cohort study of adults aged ≥18 years to <105 years residing in Ontario, Canada, with a primary cancer diagnosed between April 1^st^, 2010, and August 31^st^, 2023, who received intra-cranial RT. Patients with multiple primary cancer diagnoses before IMD diagnosis, a primary intracranial tumour diagnosis, or missing treatment data were excluded. Patients were additionally excluded if they were not eligible for the Ontario Health Insurance Plan (OHIP) six months before the index date, had an index date equal to or later than the death date, or were not Ontario residents at index.

### Dataset sources

Patients were identified using the Institute for Clinical Evaluative Sciences (IC/ES) database.^7^ IC/ES is a non-profit organization that operates per Ontario’s health information privacy law, which permits it to collect and analyze patient healthcare and demographic information without patient consent for the evaluation and improvement of our healthcare system. IC/ES houses administrative data on health outcomes of all cancer patients in Ontario, Canada. Within IC/ES, seven datasets were used: Same Discharge Abstract Database (DAD), National Ambulatory Care Reporting System (NACRS), Registered Persons Database (RPDB), Cancer Activity Level Reporting (ALR), New Drug Funding Program (NDFP), Ontario Cancer Registry (OCR), and Symptom Management Database (ESAS) (Supp. Table 1). Data was stored and analyzed following IC/ES confidentiality and security regulations. In accordance with section 45 of the Personal Health Information Protection Act (PHIPA), the project did not require patient consent and was considered exempt from research ethics board review. All datasets were linked using unique encoded identifiers and analyzed at ICES.

### Outcomes

The primary endpoint used in this study is overall survival (OS) from IMD, which was calculated as the time (in months) from the date of IMD diagnosis to death or date of last follow-up. Patient follow-up data was collected until December 31^st^, 2023. Patients who were alive at last follow-up were censored.

### Treatment & variable classifications

Patients treated with intra-cranial RT were identified using treatment codes in the ALR dataset (Supp. Table 2). RT was classified as SRT or WBRT. SRT was defined as treatment with the treatment codes of: gamma knife, cyberknife (CK) intra-cranial, linac SRT with cones, tomotherapy stereotactic body radiotherapy (SBRT) (dose/fraction ≥ 5Gy), tomotherapy (dose/fraction ≥ 5Gy), linac volumetric modulated arc therapy (VMAT) (dose/fraction ≥ 5Gy), linac intensity modulated radiation therapy (IMRT) (dose/fraction ≥ 5Gy), or Linac SBRT (dose/fraction ≥ 5Gy). WBRT was defined as treatment with the treatment codes of: Linac non-mod, Linac field-in-field, tomotherapy SBRT (dose/fraction < 5Gy), tomotherapy (dose/fraction < 5Gy), Linac VMAT (dose/fraction < 5Gy), Linac IMRT (dose/fraction < 5Gy), or Linac SBRT (dose/fraction < 5Gy) RT treatment. Patients who received multiple modalities of RT, defined as receiving a combination of SRT and WBRT techniques, were excluded.

SRT-dedicated centres were defined as those that had a dedicated intracranial apparatus (Gamma Knife or CyberKnife) and on-site neurosurgical services. Non-SRT-dedicated centres were defined as those that utilized standard image-guided multi-leaf collimator-equipped linear accelerator (LINAC) technology for intracranial RT, with or without on-site neurosurgical services.

Systemic treatment was identified using NDFP. Treatment types were categorized into chemotherapies, antibodies, antibody conjugates, immunomodulators, systemic radiation, and other treatments (Supp. Table 3). Binary values were assigned for each treatment type.

Primary cancer diagnosis was identified using OCR through either morphology or topography codes. Grouping by primary tumour type was: colon/GI; melanoma, renal cell, thyroid, sarcomas (grouped as radioresistant); lung; other; breast, based on historical radioresistance groupings and expert consultation (Supp. Table 4).^8,9^

Clinical variables identified included best stage at diagnosis, age at diagnosis, RT treatment body region code, RT intent, systemic therapy drug used, lesion counts, and symptom management scores based on the Edmonton Symptom Assessment System (ESAS) which includes criteria for anxiety, depression, drowsiness, lack of appetite, nausea, pain, shortness of breath, tiredness, wellbeing, constipation, diarrhea, sleep, and activities and function. Patients with missing data for their ESAS score were still included in Cox regression analyses. IMD lesion number was inferred from the number of SRT treatment inputs received in one session; lesion counts could only be determined within the SRT sub-cohort. Following IC/ES regulations, blinding of lesion counts within the non-SRT-dedicated program centres subgroup was implemented.

Location variables identified included the centre at which systemic treatment and RT were delivered. Discrepancies in the treatment centre name were consolidated by grouping centres for which the centre name was different but where centres fell under the auspices of a single cancer centre unit. Centres were categorized as systemic treatment-only centres, non-SRT-dedicated program centres, and SRT-dedicated program centres.

### Statistical Analysis

Group differences in patient characteristics were compared using two-sample t-tests without assumed equal variances (for continuous variables), odds ratios (for binomial categorical variables), and chi-squared tests (for non-binomial categorical variables). Patients were grouped in two ways, comprising two sets of analyses: 1) patients who received RT treatment at an SRT-dedicated vs. an non-SRT-dedicated centre; and 2) patients who moved centres vs. those who stayed at the same centre at which their systemic treatment was administered. A third analysis was performed by stratifying patients based on movement between SRT-dedicated program sites and non-SRT-dedicated program sites over the course of systemic and RT treatment and comparing mean OS using a t-test without assumed equal variances. Odds ratios performed where one of the categories has a value of zero were corrected using a continuity correction of 0.5.

Kaplan-Meier analysis was performed to estimate and visualize OS. Kaplan-Meier and Cox-proportional hazard regression models were used to calculate hazard ratios (HRs) for OS. In Cox regression models, covariates included systemic treatment, age at primary diagnosis, ESAS score, sex, and primary tumour type. Two comparative analyses for OS were performed. First, the HR for OS was compared between patients receiving RT at a SRT-dedicated vs. non-SRT-dedicated program centres. Second, OS was compared based on patient movement between receipt of systemic therapies and RT treatment, testing the impact of movement to or from SRT-dedicated or non-SRT-dedicated centres. Bonferroni p-value correction was applied to account for multiple testing bias, with a corrected two-sided p-value < 0.05 considered statistically significant. For all tests, uncorrected models (i.e. those without covariates) are first presented, followed by multivariate Cox regression analyses to determine sensitivity; therefore, p-value corrections for multiple testing were not applied to Cox regression models.

## Results

### Patient characteristics across centres with or without dedicated SRT programs

The patient cohort included 4,410 patients, 54% (n = 2370) of whom received RT treatment at an SRT-dedicated centre and 46% (n = 2040) at a non-SRT-dedicated centre. The mean number of lesions treated with SRT within the whole cohort was 2.1 (SD 2.1-2.2), 2.6 (SD 2.4-2.7) in patients treated at an SRT-dedicated centre and 1.5 (SD 1.4-1.5) in patients treated at a non-SRT-dedicated centre (p < 0.0001). Between centre types, the ratio of 4 or less vs. 5 or more lesions treated with SRT showed a statistically significant difference (OR = 29.9, 95% CI 14.5 − 76.0, p < 0.0001). More patients with higher lesion counts were treated with SRT at SRT-dedicated centres than at non-SRT-dedicated centres (Table 1).

**Table 1.**
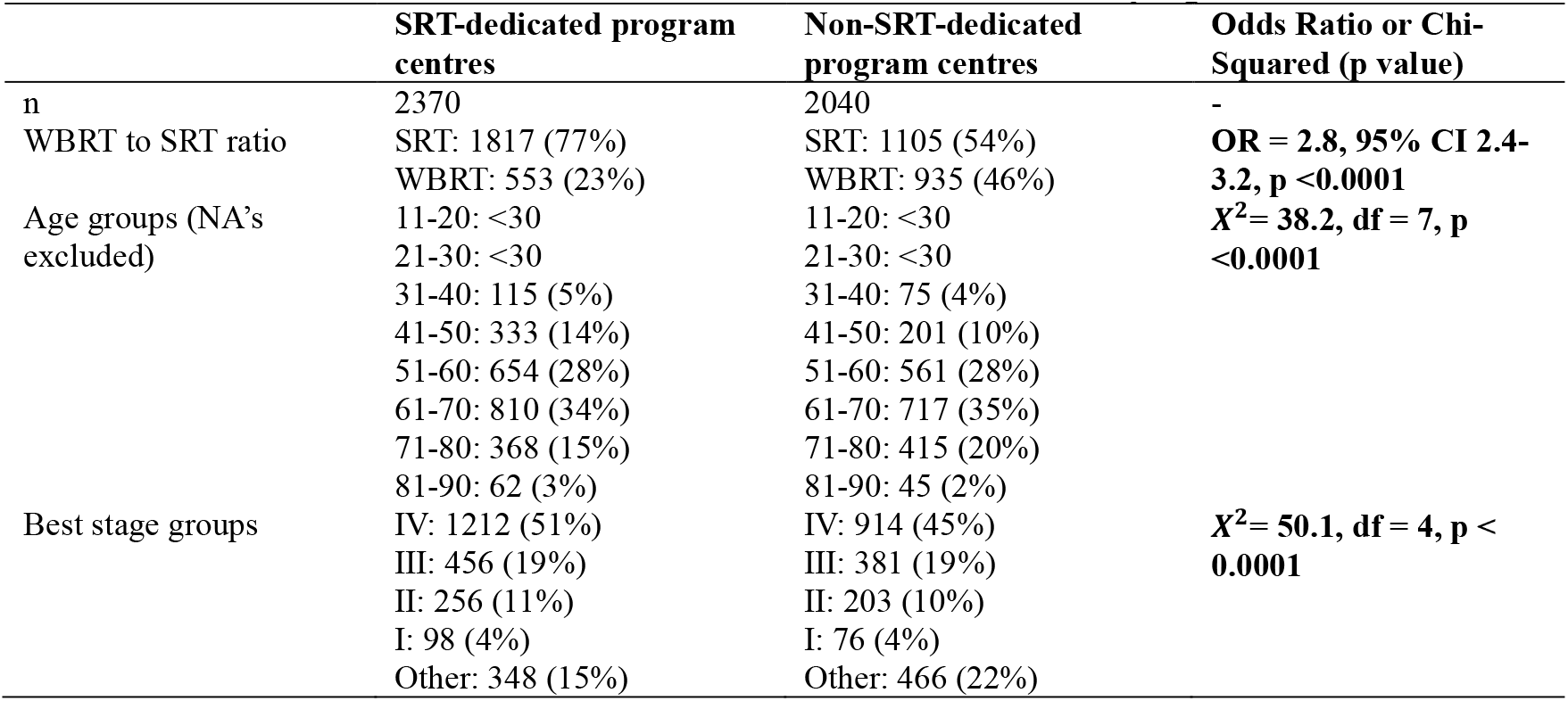
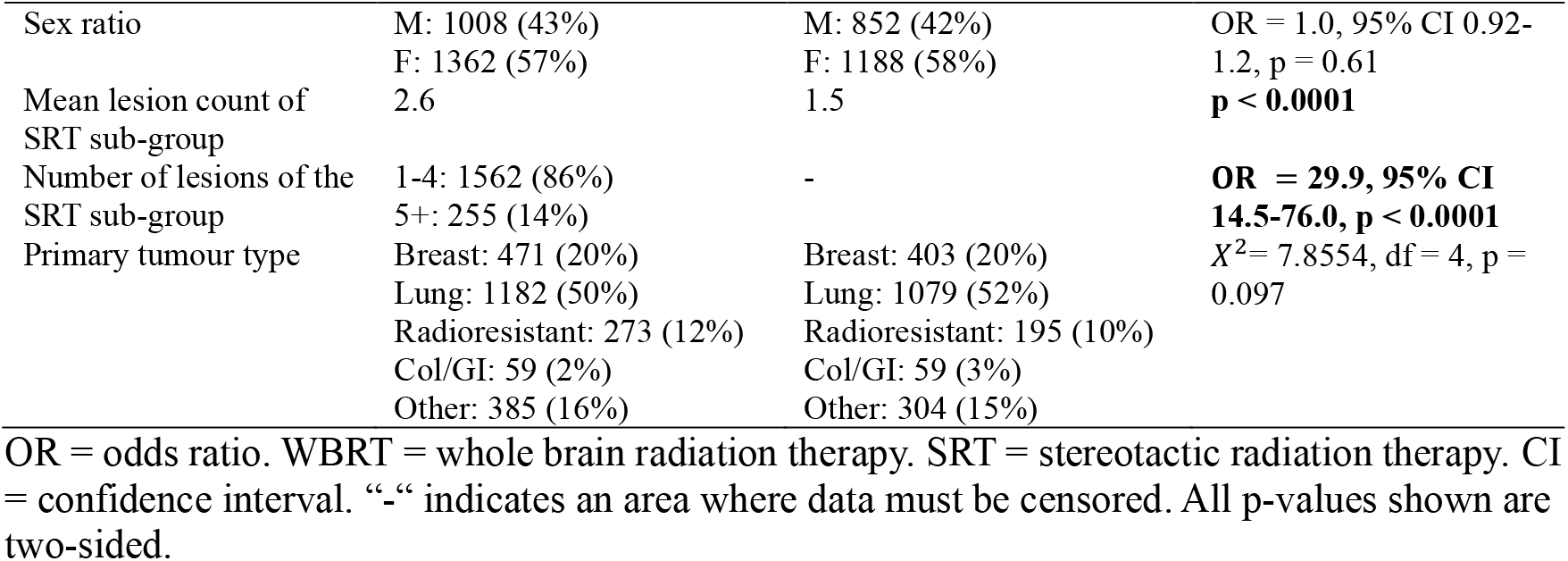
Patient characteristics at centres with/without dedicated SRT programs.

We then sought to test our hypothesis of differential SRT and WBRT usage across SRT-dedicated and non-SRT-dedicated centres. We found that more patients received SRT relative to WBRT at SRT-dedicated vs non-SRT-dedicated centres (OR = 2.8, 95% CI 2.4-3.2, p <0.0001). Patient age at treatment was lower at SRT-dedicated centres (mean 60.1 years) than at non-SRT-dedicated centres (mean 61.8 years); non-SRT-dedicated program centres tended to treat older patients (t = 4.8, p <0.0001), with non-SRT-dedicated program centres treating a higher proportion of patients aged 71-80 (*X*^2^ = 38.2, df = 7, p < 0.0001, Table 1). A higher proportion of patients at SRT-dedicated centres were stage IV in their best stage at initial cancer diagnosis, relative to non-SRT-dedicated centres (*X*^2^=50.1, df = 4, p < 0.0001, Table 1). Between RT centre types, there were no significant differences in sex or radioresistant/radiosensitive primary tumour type (i.e. breast, lung, radioresistant, colon/GI, or other) (Table 1).

### Overall survival for SRT and WBRT across treatment centre categories

In bivariate Kaplan-Meier models, the median OS for patients treated with SRT vs WBRT in an SRT-dedicated centre was 10.15 vs 2.96 months, respectively (HR = 0.44, 95% CI = 0.40-0.49, p < 0.0001). Within non-SRT-dedicated centres, median OS in patients treated with SRT vs. WBRT was 7.89 vs. 3.09 months, respectively (HR = 0.55, 95% CI = 0.50-0.61, p < 0.0001). These differences remained significant after controlling for covariates using Cox regression (p < 0.0001).

Comparing patients treated with SRT at an SRT-dedicated vs. non-SRT-dedicated centre, median OS was 10.15 vs 7.89 months (HR = 0.88, 95% CI, 0.81-0.96, p = 0.028), respectively. These results remained significant after controlling for covariates (p < 0.001; Table 2, Fig. 1A). The median OS of patients treated with WBRT at either an SRT-dedicated vs. non-SRT-dedicated centre was 2.96 vs 3.09 months (HR = 1.08, 95% CI, 0.97-1.20, p = 1.0), respectively. This negative finding was consistent with Cox regression modelling (p = 0.38; Table 2, Fig. 1B).

**Table 2.** Cox-proportional hazards for overall survival of patients receiving radiation therapy treatment at either SRT-dedicated or non-SRT-dedicated centres.

| Centre type | Median OS (months) | Kaplan-Meier analysis HR | Adjusted P value (Bonferroni) | Cox regression analysis HR |
| --- | --- | --- | --- | --- |
| Only SRT tx |  |  |  |  |
| SRT-centre | 10.15 (95% CI, 9.27-11.33) | 0.88 (95% CI, 0.81-0.96), p = 0.0028 | p = 0.028 | 0.83 (95% CI, 0.75-0.92), p < 0.001 |
| Non-SRT centre | 7.89 (95% CI, 7.03-8.71) |  |  |  |
| Only WBRT tx |  |  |  |  |
| SRT-centre | 2.96 (95% CI, 2.53-3.32) | 1.08 (95% CI, 0.97-1.20), p = 0.17 | p = 1.0 | 1.06 (95% CI, 0.93-1.21), p = 0.38 |
| Non-SRT centre | 2.73 (95% CI, 2.73-2.52) |  |  |  |

**Figure 1.**
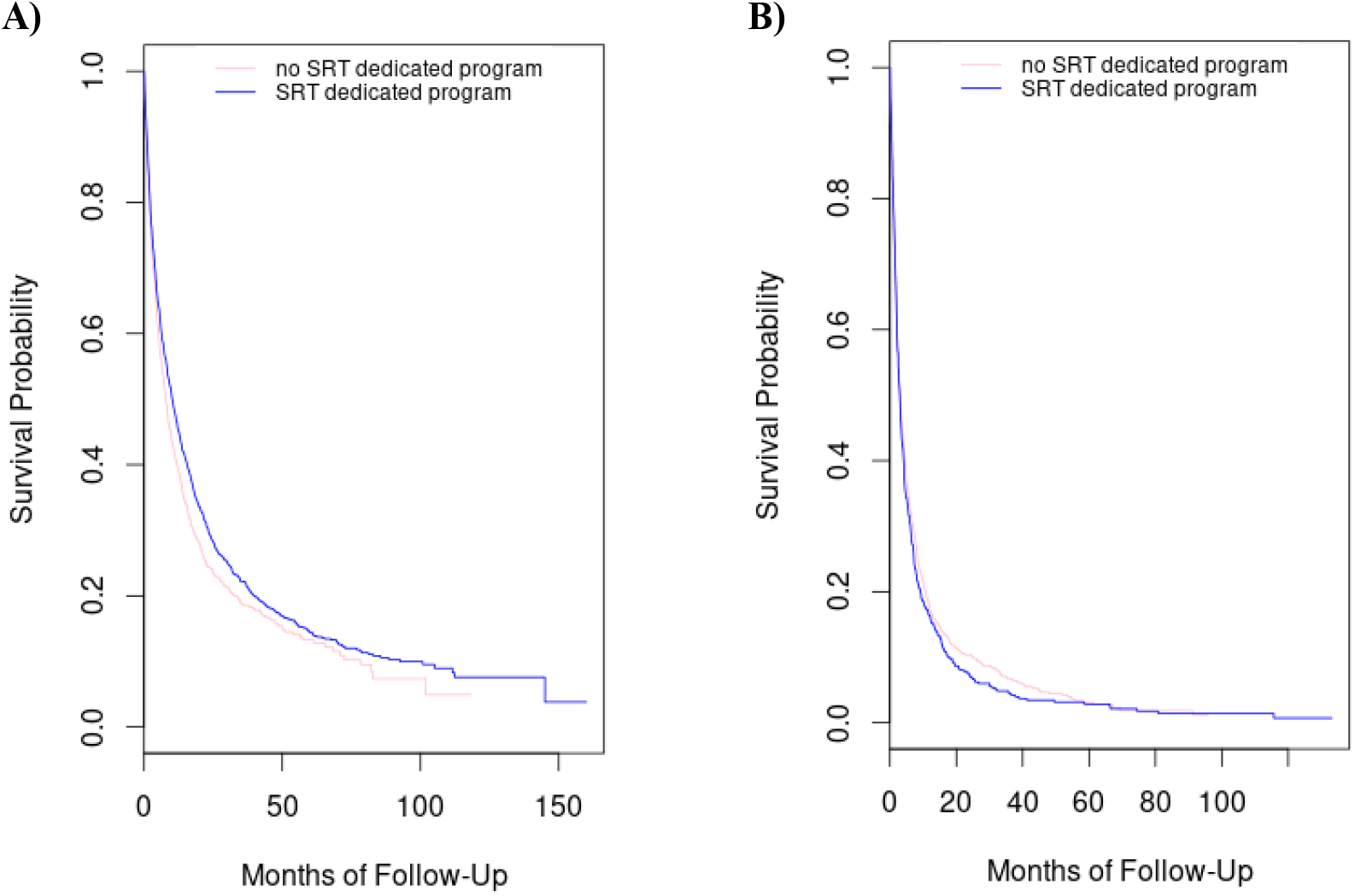
Kaplan-Meier curves of overall survival between IMD patients A) receiving SRT at either SRT or non-SRT program centres, B) receiving WBRT at either SRT or non-SRT program centres.

### Characteristics and outcomes of patients switching centres for treatment

We compared outcomes in patients who either stayed at the same centre for systemic and RT treatment vs. those who switched centres between treatments, excluding patients who received systemic treatment at a centre at which RT was not offered. No significant differences in age group, primary tumour type, or sex were found (Table 3).

**Table 3.** Centre movement between systemic and RT treatment based on patient characteristics; excluding patients who received systemic treatment at a non-RT centre or were missing location data for systemic or RT treatment.

|  | Same centre for systemic and RT treatment | Different centre for systemic and RT treatment | Odds Ratio or Chi-Squared (p-value) |
| --- | --- | --- | --- |
| n | 1793 | 304 |  |
| age group | 11-20: <25 | 11-20: <25 | $X^2=6.01$, df = 7, p = 0.54 |
|  | 21-30: <25 | 21-30: <25 |  |
|  | 31-40: 89 | 31-40: 23 |  |
|  | 41-50: 250 | 41-50: 42 |  |
|  | 51-60: 500 | 51-60: 77 |  |
|  | 61-70: 580 | 61-70: 101 |  |
|  | 71-80: 322 | 71-80: 51 |  |
|  | 81-90: 31 | 81-90: <10 |  |
| primary tumour category | Breast: 459 | Breast: 81 | $X^2=8.06$, df = 4, p = 0.090 |
|  | Lung: 722 | Lung: 136 |  |
|  | Radioresistant: 204 | Radioresistant: 38 |  |
|  | Colon/GI: 43 | Colon/GI: 8 |  |
|  | Other: 365 | Other: 41 |  |
| Sex | M: 700 | M: 113 | OR = 0.92, 95% CI 0.72-1.2, p = 0.54 |
|  | F: 1093 | F: 191 |  |

OR = odds ratio.

Of the 800 patients who received systemic treatment at a non-RT centre, 382 received RT at an SRT-dedicated centre (received SRT: n = 265; received WBRT: n = 117; OR = 1.6, 95% CI 1.2-2.1) vs. 418 at a non-SRT-dedicated centre (received SRT: n = 245; received WBRT: n = 173) (p = 0.0016). Patients who received systemic treatment at a non-SRT-dedicated centre were more likely to move to a different centre for their intracranial RT treatment than those who received systemic treatment at an SRT-dedicated centre (OR = 3.9, 95% CI 3.0-5.3, p < 0.0001). Patients who received systemic treatment at a non-SRT-dedicated centre and required intracranial SRT were more likely to move centres between systemic and RT treatment (OR = 7.6, 95% CI 5.2-11.5, p < 0.0001, Fig. 2A), compared to patients receiving systemic treatment at an SRT-dedicated centre (OR = 0.38, 95% CI 0.22-0.63, p < 0.001, Fig. 2A).

**Figure 2.**
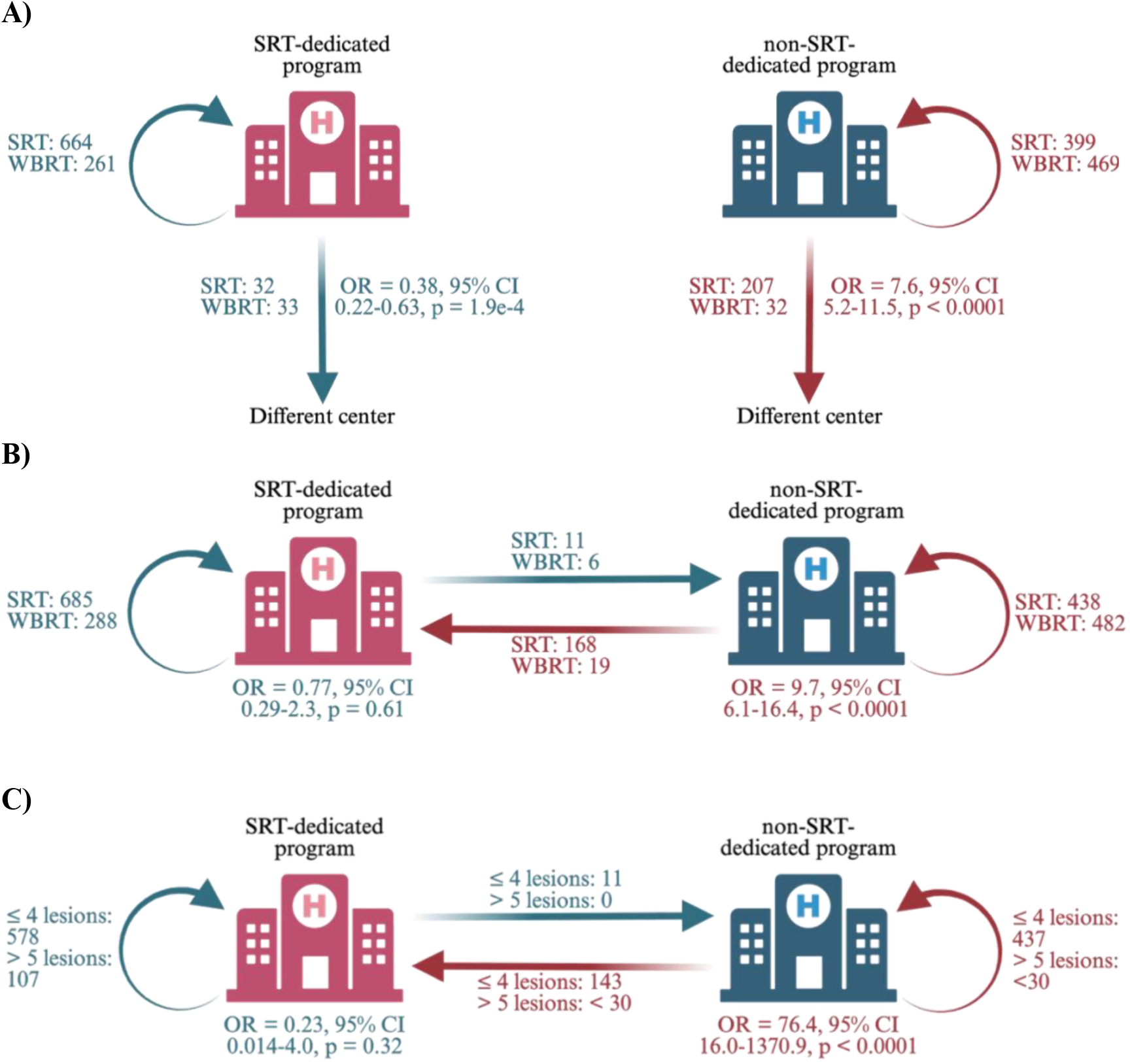
An illustration describing patient movement between receipt of systemic and RT treatments. A) Patients who started at and stayed at an SRT-dedicated program centre vs patients who moved to a different centre based on receipt of SRT or WBRT (OR = 0.38, 95% CI 0.22-0.63, p = 1.9e-4); and patients who started at and stayed at a non-SRT-dedicated program centre vs patients who moved to a different centre based on receipt of SRT or WBRT (OR = 7.6, 95% CI 5.2-11.5, p < 0.0001). B) Patients who stayed at an SRT-dedicated program centre vs patients who moved to a non-SRT-dedicated program centre based on receipt of SRT or WBRT (OR = 0.77, 95% CI 0.29-2.3, p = 0.61); and patients who stayed at a non-SRT-dedicated program centre vs patients who moved to an SRT-dedicated program centre based on receipt of SRT or WBRT (OR = 9.7, 95% CI 6.1-16.4, p < 0.0001). C) Patients who stayed at an SRT-dedicated program centre vs patients who moved to a non-SRT-dedicated program centre based on lesion counts when treated with SRT (OR = 0.23, 95% CI 0.014-4.0, p = 0.32); and patients who stayed at a non-SRT-dedicated program centre vs patients who moved to an SRT-dedicated program centre based on lesion counts when treated with SRT (OR = 76.4, 95% CI 16.0-1370.9, p < 0.0001).

Although patient movement in various directions was observed, movement from one RT-capable centre to another was most likely in patients moving from a non-SRT-dedicated centre to an SRT-dedicated centre for SRT (OR = 9.7, 95% CI 6.1-16.4, p < 0.0001, Fig. 2B), relative to patients moving from an SRT-dedicated centre to a non-SRT-dedicated centre for SRT (OR = 0.77, 95% CI 0.29-2.3, p = 0.61, Fig. 2B). Patients with higher lesion counts who moved centres for SRT most frequently moved from a non-SRT-dedicated centre to an SRT-dedicated centre (OR = 76.4, 95% CI 16.0-1370.9, p < 0.0001, Fig. 2C); all patients who moved from an SRT-dedicated to a non-SRT-dedicated centre for SRT had four or fewer lesions (OR = 0.23, 95% CI 0.014-4.0, p = 0.32, Fig. 2C).

On t-test analysis, a positive association in mean OS was observed in patients who received systemic treatment at an non-SRT-dedicated centre and moved to an SRT-dedicated centre for RT treatment (14.0 months) compared to the mean OS of patients who received both treatments at non-SRT-dedicated centres (7.3 months, t = -3.90, df = 209, p < 0.001).

## Discussion

This provincial-level, administrative database-based, retrospective cohort study investigated the outcomes of patients with IMD who received SRT or WBRT in Ontario, Canada, in an centre-specific manner. SRT-dedicated program centres used SRT more frequently, and in patients with greater lesions counts, than did non-SRT-dedicated program centres. Prolonged OS was observed in patients treated with SRT at SRT-dedicated centres, compared to patients treated with SRT at non-SRT-dedicated centres. We surmise that this observation is linked to hardware and clinician factors that differentiate SRT-dedicated and non-SRT-dedicated centers, such as the presence of a dedicated intracranial SRT system and central nervous system-specialized multidisciplinary teams, including radiation oncology, medical physics, neuroradiologists, neuro-oncologists, and neurosurgeons, that allow for both rapid case discussion and surgical management of complications and tumour progression. Although the difference in OS rates were significant, we acknowledge that the 2.8 month absolute difference is modest.

We did observe differences across the two centre types in age groups, best stage at diagnosis, and number of lesions treated with SRT. A higher proportion of older patients were treated at non-SRT-dedicated centres, which likely reflects more judicious allocation of resources at non-SRT centers with the aim to avoid WBRT in the elderly. Furthermore, we surmise that older patients are likely less able and motivated to travel long distances for their IMD care. Conversely, a higher proportion of patients with more advanced stage at diagnosis were treated at SRT-dedicated centres, which likely reflects the ability of centres to be more liberal in their indications to treat IMD, given the presence of a dedicated brain SRT apparatus. Similarities were also observed between SRT and non-SRT centers outcomes, in particular equivalent OS outcomes following WBRT. This observation indirectly provides additional support to the conclusion that the SRT component of care in a SRT-dedicated center accounts for the longer patient survival seen with treatment at SRT-dedicated centres.

When comparing OS for patients who stayed at a non-SRT-dedicated centre for systemic and RT treatment with OS for patients who moved to an SRT-dedicated centre following systemic therapy, a significant difference was observed favoring the latter.

While we were able to assess a large cohort of patients and compare geographic treatment and patient outcome trends within Ontario, our study has several intrinsic limitations. First, this was a retrospective study that utilized an administrative database and, as such, we were unable to consider issues regarding treatment planning and treatment decision-making. For example, assuming that the centre at which patients received systemic treatment was likely the closest/most available centre to them, we were unable to discern whether a decision to undertake RT at that centre reflected a determination by the local physician that the treatment plan offered was adequate, if treatment and movement were due to a referee bias incurred by physicians, or if the patient requested local treatment despite recommendations for treatment at an SRT-dedicated centre. Second, our access to performance status data and information on extracranial disease burden at the time of cranial RT were limited; we were thus unable to critically assess the context of patient referral to SRT-dedicated or non-SRT-dedicated centres aside from factors such as type of RT treatment received and lesion counts in patients receiving SRT. Third, the data did not allow us to determine lesion count in patients who were treated with WBRT. Finally, without access to data directly related to the cancer and its treatments, we were unable to assess any incurred patient complications.

## Conclusion

In this large, retrospective cohort study, we found greater utilization of SRT and longer OS in patients who were treated in centres with SRT-dedicated programs. OS was positively impacted for patients who travelled to a centre with a dedicated SRT program for radiation therapy.

## Data Availability

All data produced in the present study are kept by the Institute for Clinical Evaluative Sciences

## Acknowledgments

This study was supported by ICES. Part of this material are based on data provided and compiled by Ontario Health (OH), CIHI, and the Ontario Ministry of Health (MOH). The analyses, opinions, statements, and conclusions expressed are solely those of the authors and do not reflect those of the funding or data sources. No endorsement is intended or should be inferred. Figure 2 was created in BioRender.

