## Supplemental tables 1-4 for "Centre-specific treatment and outcomes of patients with brain metastases treated with whole brain or stereotactic radiation therapy: an Ontario-based retrospective cohort study"

**Supplemental Materials**

**Supplemental Table 1.** Categorization of data types based on dataset origin.

| **Data type** | **Origin dataset and resolution if applicable** |
| --- | --- |
| Sex | RPDB |
| Death date | RPDB |
| ICD-0-3 topography code | OCR |
| ICD-0-3 morphology code | OCR |
| Best stage at diagnosis (collaborative stage summary stage or the pathological/clinical stage from case resolution) | OCR |
| Diagnosis age | OCR |
| Date of primary cancer diagnosis | ALR |
| IMD flag | ALR, DAD, NACRS |
| IMD date | ALR, DAD, NACRS; if inconsistent IMD date, resolved by using the dates ALR > DAD and SDS > NACRS |
| Radiation tx date | ALR |
| Radiation tx body region code | ALR |
| Course of radiation treatment | ALR |
| Intent of radiation treatment | ALR |
| NHPIP code | ALR |
| Systemic therapy tx date | NDFP |
| Drug name | NDFP |
| Description (description of eligible disease and indication) | NDFP |
| ESAS – symptom management results | ESAS |
| Radiation treatment location data | ALR |
| Systemic treatment location data | NDFP |

**Supplemental Table 2.** NHPIP external beam treatment delivery codes. Bolded treatments are the defined relevant treatments.

| **500** | Tx Superficial X-Ray |
| --- | --- |
| **510** | Tx Orthovoltage |
| **538** | Tx Linac TBI |
| **540** | Tx Linac Electrons |
| **542** | Tx Linac TSE |
| **543** | **Tx Linac Non-Mod** |
| **544** | **Tx Linac Fld-in-Fld** |
| **548** | **Tx Linac SRS w Cones** |
| **592** | **Tx Linac IMRT** |
| **593** | **Tx Linac SBRT** |
| **594** | **Tx Linac VMAT** |
| **572** | **Tx Gammaknife** |
| **598** | **Tx CK Intra-Cranial** |
| **599** | Tx CK Extra-Cranial |
| **575** | **Tx Tomotherapy** |
| **576** | Tx Tomotherapy TMI |
| **595** | **Tx Tomotherapy SBRT** |
| **582** | Tx PDT |
| **590** | Tx Radionuclide |
| **701** | Internal Eye Shield |

**Supplemental Table 3.** Systemic treatment categorization.

| **Treatment category** | **Treatment** |
| --- | --- |
| Chemotherapy | Paclitaxel, Pemetrexed, Docetaxel, Bendamustine, Azacitidine, Cavazitaxel, Epirubicin, Irinotecan, Oxaliplatin, Gemcitabine, Vinorelbine, Topotecan, Bab-Paclitaxel, Eribulin, Liposomal Doxorubicin, Raltitrexed, Pegaspargase, Pralatrexate, Fludarabine, Liposomal daunorubicin and Cytarabine, Arsenic Trioxide, Thiotepa |
| Antibody | Nivolumab, Ipilimumab, Pembrolizumab, Rituximab IV (Rituxan), Trastuzumab (Herceptin), Pertuzumab, Rituximab IV (Truxima), Rituximab SC, Bevacizumab (Mvasi), Atezolizumab, Bevacizumab (Avastin), Ramucirumab, Bevacizumab (Zirabev), Trastuzumab (Trazimera), Panitumab, Denosumab, Daratumumab SC, Obinutuzumab, Trastuzumab (Ogivri), Daratumumab IV, Rituximab IV (Riximyo), Rituximab IV (Ruxience), Durvalumab, Avelumab, Trastuzumab (Kanjinti), Cetuximab, Blinatumomab_overfill, Blinatumomab, Trastuzumab (Herzuma), Cemiplimab, Bevacizumab (Bamvevi), Istatuximab, Siltuximab, Dinutuximab |
| Antibody conjugate | Trastuzumab emtansine, Polatuzumab vedotin, Brentuximab vedotin, Gemtuzumab ozogamicin, Trastuzumab deruxtecan, enfortumav vedotin, Sacituzumav govitecan, Inotuzumab Ozogamicin |
| Immunomodulator | Interferon, Aldesleukin, Interferon lyophilized powder |
| Systemic radiation | Radium-223 dichloride, Strontium-89 |
| Other | Pamidronate, Carfilzomib, Bortezomib, Zoledronic acid, Plerixafor, Midostaurin, Ventoclax, Temsirolimus, Romidepsin, Gilteritinib, Porfimer sodium, Erwinia asparaginase |

**Supplemental Table 4.** Primary cancer morphology and topography codes.

| **Topography** |  |
| --- | --- |
| Gastro-intestinal | C260-269 |
| Melanoma | C440-449, C510-519, C600-609, C632 |
| Thyroid | C739 |
| Lung | C340-349 |
| Breast | C500-C509 |
| Colon | C180-189 |
| **Morphology** |  |
| Renal cell | 83123, 83163, 83173, 83183, 82603 |
| Sarcoma | 86800-87139, 8800-89219, 89900-89919, 90400-90449, 91200-91259, 91300-91369, 91410-92529, 93700-93739, 95400-95829 |
